# A Novel Use of Intracranial Arterial Pressure Waveforms to Detect Occlusive events

**DOI:** 10.1101/2025.04.28.25326342

**Authors:** Ryuzaburo Kochi, Eishi Asano, Yoshiteru Shimoda, Atsushi Kanoke, Shunsuke Omodaka, Hiroyuki Sakata, Kanako Sato, Yasuhiro Suzuki, Kuniyasu Niizuma, Yasushi Matsumoto, Hidenori Endo

## Abstract

During neuroendovascular treatment, unexpected occlusive events, such as thrombosis, may occur and can cause severe complications. However, angiography provides only intermittent assessments of intravascular conditions, underscoring the need for continuous monitoring. In this retrospective study, we examined whether intracranial arterial pressure waveforms recorded via a catheter could detect occlusive events. A total of 6,442 waveform trials from 43 arteries in 37 patients, each assessed in both patent and occluded states, were analyzed. Time-frequency amplitude (TFA) was calculated via wavelet transformation in selected frequency ranges (6 to 10, 10 to 14, 14 to 18, and 6 to 18 Hz). A mixed-model analysis revealed that TFA was significantly higher under occluded conditions (p < 0.001). Specifically, when focusing on the 6 to 18 Hz range and using a 113.9% elevation in TFA from the initial patent baseline as a threshold, the average sensitivity, specificity, positive predictive value, and negative predictive value for detecting occlusion were 88.5%, 87.5%, 36.1%, and 96.4%, respectively. These findings suggest that continuous waveform analysis may serve as a valuable complement to intermittent angiographic assessments, potentially enhancing both safety and therapeutic outcomes in neuroendovascular procedures.

## Introduction

Unexpected occlusive events during neuroendovascular treatment, such as thrombosis, can result in severe complications^1-3^. Early detection is crucial because it allows prompt intervention to minimize adverse outcomes^4-7^. Although angiography remains the standard for evaluating intravascular states, it only provides information during contrast administration and radiation exposure, leading to intermittent observation. As a result, occlusions may go undetected if they occur between imaging sessions. In addition, repeated angiography is frequently required to monitor rapidly changing intravascular states, for example transitions between occluded and patent states following thrombotic events, which increases both radiation exposure and contrast media use. Therefore, a continuous monitoring modality that offers current intravascular state information is highly desirable. Moreover, using such a modality during intentional occlusive procedures such as parent artery occlusion with coil or intravascular balloon expansion could reduce radiation dose and contrast use by providing current intravascular status without extra imaging.

Intracranial arterial pressure (IAP) waveforms can be continuously measured through a catheter inserted during neuroendovascular treatment. Characteristic alterations in blood pressure waveforms are known to reflect intravascular hemodynamic states, as they are thought to vary in response to factors such as peripheral resistance and stroke volume^8-10^. Occlusive events may cause drastic changes in these factors, thereby altering the waveforms. Such alterations can be captured quantitatively through time-frequency analysis^11, 12^. Therefore, we hypothesize that time-frequency analysis of waveforms can differentiate between occluded and patent states.

In this study, we investigated whether quantifiable features derived from time-frequency analysis of waveforms can reflect intravascular states by comparing features obtained under occluded and patent states. Furthermore, we aimed to develop a predictive tool for detecting occluded states using cutoff values.

## Methods

### Study Inclusion

We performed a retrospective analysis of consecutive patients who underwent neuroendovascular treatment at Kohnan Hospital from February to March 2022 and at Iwaki Medical Center from April to December 2022.

### Data Acquisition and Preprocessing

IAP was continuously recorded using a pressure monitoring kit (Merit Medical Systems, Utah, USA) connected to a guiding or angiographic catheter during each procedure. Periods containing artifacts caused by manual manipulations of the catheter were removed. The remaining data were then segmented and labeled as either patent or occluded based on angiographic findings. Specifically, segments recorded between two angiographic assessments confirming patency were labeled as patent, whereas segments were labeled as occluded if they were recorded between two assessments confirming occlusion, or during the period following complete balloon inflation and before deflation.

Each segment was processed with a 1–40 Hz bandpass filter and subdivided into individual waveform trials corresponding to each RR interval derived from the concurrently recorded electrocardiogram (ECG). R peaks were detected in each 5-second interval following bandpass filtering of the ECG signal at 10–40 Hz. MATLAB’s findpeaks function was then used with MinPeakHeight = 0.2, MinPeakProminence = 0.1, and MinPeakDistance = 0.15 seconds. RR intervals with ectopic beats that failed to produce a corresponding IAP waveform were excluded. Time-frequency analysis was then applied to each waveform by performing a continuous wavelet transform (CWT) across the entire RR interval to obtain CWT amplitude values. For each waveform, an integration window was defined from the first bottom peak of its fourth derivative to 30% of the waveform’s total duration. This window encompasses the second zero-crossing point of the fourth derivative, which marks the onset of the backward wave^13, 14^. Within this time window, we integrated the CWT amplitude values across 4 frequency bands (6–10 Hz, 10–14 Hz, 14–18 Hz, and 6-18 Hz). Frequencies below 6 Hz were excluded because they are largely influenced by the low-frequency component, which is the primary component of the waveform. The integrated CWT amplitude values from these bands were used as dependent variables in all subsequent analyses, hereafter referred to as time-frequency amplitude (TFA).

### Mixed-Model Analysis of TFA

A mixed model analysis was conducted to assess the effects of several fixed factors on the integrated TFA for each frequency band The fixed factors included age, subarachnoid hemorrhage (SAH) status (i.e., whether the patient had SAH), occlusion states (i.e., whether the artery was occluded), waveform trial order within each segment, and the effective luminal area of the catheter connected to the pressure monitoring kit. The effective luminal area was defined as the cross-sectional area of the catheter’s lumen minus the outer cross-sectional area of the coaxial catheter contained within it. To account for interindividual variability, both the sampled artery and the intercept were included as random effects. Statistical significance was set at a Bonferroni corrected p value of < 0.05. We included SAH status as a factor because it can significantly alter intracranial dynamics, in particular by raising intracranial pressure and reducing cerebral perfusion, which may influence TFA^15, 16^. We also included the effective luminal area as a factor, because changes in this area can influence the transmission of the pressure waveform^17, 18^.

### Detection Performance Analysis at artery level

To allow comparisons across the sampled arteries, we calculated the percent change in TFA for each waveform trial. Specifically, in the first patent-state segment, the median TFA of the initial 5 waveform trials served as the reference. The percent change relative to this reference was then computed for each subsequent trial beginning with the sixth in the same segment, as well as for all trials in later segments. Whenever the effective luminal area of the catheter connected to the pressure-monitoring kit changed (e.g., due to the insertion or removal of a coaxial catheter), we recalculated the reference based on the median TFA of the first 5 waveform trials thereafter. All subsequent trials were then expressed as a percent change relative to this new reference.

A leave-one-out (LOO) analysis was then performed using the percent change in TFA. For each artery, its data was held out as the test set, while data from all other arteries were used as the training set. Within each training set, the optimal cutoff value that maximized the Youden index was identified. This cutoff was then applied to the test set, and standard classification metrics were recorded: the area under the receiver operating characteristic curve (AUROC), the area under the precision–recall curve (AUPRC), the Youden index, the best cutoff value, sensitivity, specificity, positive predictive value (PPV), and negative predictive value (NPV). Subsequently, a one-sample t test was conducted for each metric to obtain its mean, 95% confidence interval, minimum, maximum, and p value. All analyses were performed using MATLAB (R2024a, MathWorks, Natick, MA, USA) and IBM SPSS Statistics (version 30, Armonk, NY, USA).

## Results

From the recordings, a total of 6,457 RR intervals were detected from 124 segments (88 patent, 36 occluded) obtained from 43 arteries in 37 patients. Fifteen RR intervals with ectopic beats that failed to produce a corresponding IAP waveform were excluded, leaving 6,442 waveform trials for analysis. The median patient age was 62 years (range: 40–81), 23 were female and 19 were SAH patients (see Supplemental Table 1 and Supplemental Table2). An illustrative case of a patient who underwent balloon-assisted coil embolization for an unruptured intracranial aneurysm is shown in Figure 1.

**Figure 1.**
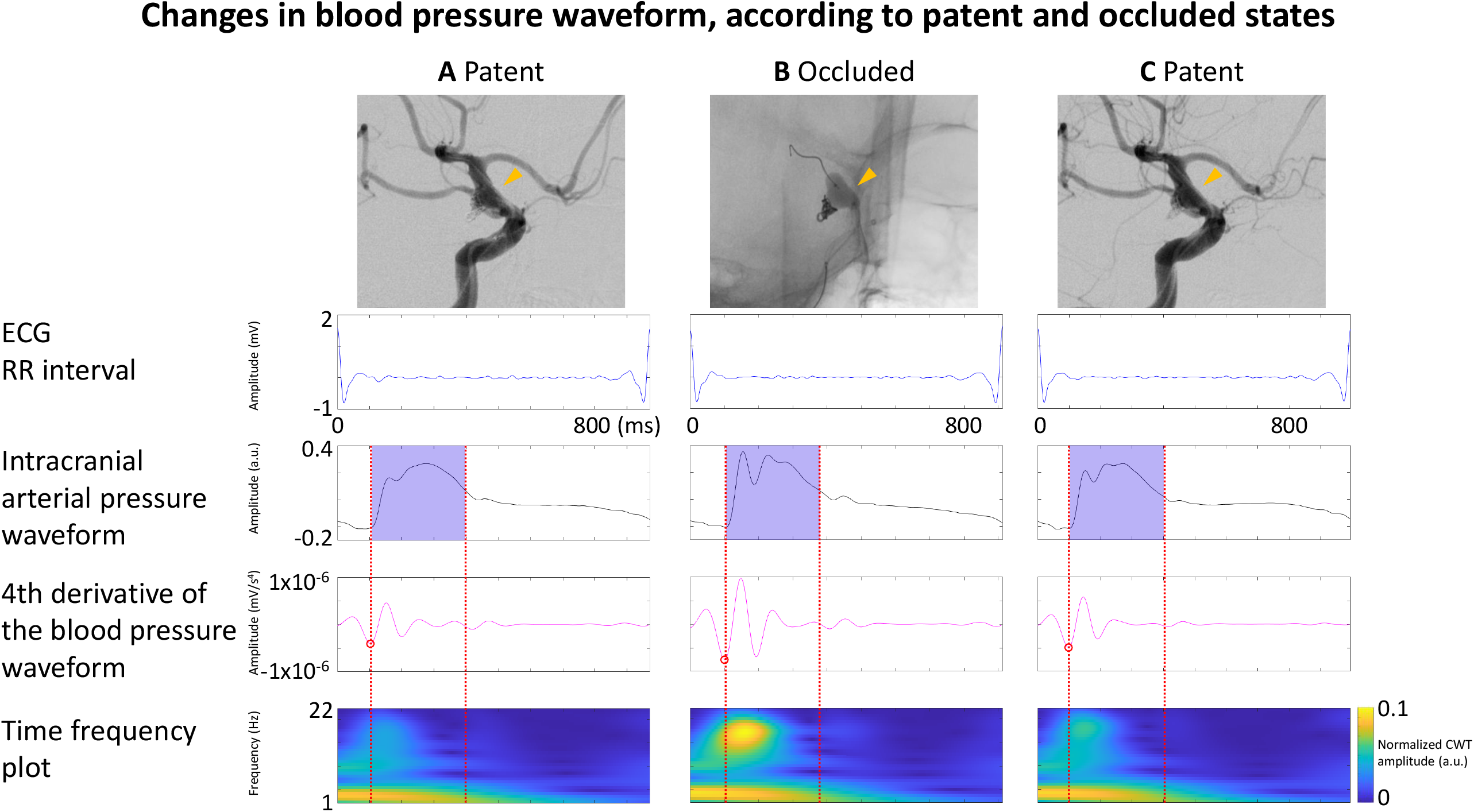
Illustrative Case (Patient 05): Treatment of balloon-assisted coil embolization for an unruptured left intracranial carotid artery aneurysm. (A) Just before balloon inflation, showing patent state. (B) After complete balloon inflation, demonstrating occluded state. (C) After complete balloon deflation, returning to patent state. The yellow arrowhead indicates the corresponding location in each angiographic image and the midpoint of the balloon. The purple area on the IAP waveforms and red broken lines represents the time window used for calculating the integrated CWT amplitude value. The red circle on the fourth derivative of the IAP waveform marks its first bottom peak. A clear augmentation of the CWT amplitude was observed in the time-frequency plot generated from the IAP waveform during occluded states (B, bottom). a.u., arbitrary unit; CWT, continuous wavelet transform; ECG, electrocardiogram. IAP: intracranial arterial pressure

### Mixed-Model Analysis of TFA

A mixed-model analysis (see Supplemental Table 3 for details) showed that TFA were positively correlated with occluded states in all four frequency bands (p < 0.001; average t-value: 45.28). They were also positively correlated with waveform trial number in all four bands (p < 0.001; average t-value: 5.02), and the effective luminal area of the catheter connected to the pressure-monitoring kit was positively correlated in all 4 bands (p < 0.001; average t-value: 34.44). All pairwise correlation coefficients were below 0.7, indicating no substantial multicollinearity. After Bonferroni correction (α = 0.05/4 ≈ 0.013), all initially significant correlations remained significant. Neither age nor SAH status showed any significant effect on TFA.

### Detection Performance Analysis at artery level

The LOO analysis and subsequent one-sample t tests yielded the following findings. In the 6–18 Hz band, the mean AUROC, AUPRC, best cutoff, sensitivity, specificity, PPV, and NPV were 88.1% (p < 0.001; 95% CI: 87.8–88.4), 77.2% (p < 0.001; 95% CI: 76.8–77.5), 113.9 (p < 0.001; 95% CI: 113.8–114.1), 88.5% (p < 0.001; 95% CI: 74.3–102.8), 87.5% (p < 0.001; 95% CI: 81.2–93.8), 36.1% (p < 0.001; 95% CI: 19.7–52.4), and 96.4% (p < 0.001; 95% CI: 93.0–99.8), respectively. In the 6–10 Hz band, the respective means were 83.9% (p < 0.001; 95% CI: 83.6– 84.2), 70.6% (p < 0.001; 95% CI: 70.3–70.9), 107.4 (p < 0.001; 95% CI: 107.2–107.7), 75.9% (p < 0.001; 95% CI: 55.0–96.8), 79.1% (p < 0.001; 95% CI: 71.7–86.5), 29.6% (p < 0.001; 95% CI: 15.7–43.5), and 94.1% (p < 0.001; 95% CI: 89.0–99.2). In the 10–14 Hz band, the mean values were 83.9% (p < 0.001; 95% CI: 83.6–84.2), 75.9% (p < 0.001; 95% CI: 75.5–76.4), 114.1 (p < 0.001; 95% CI: 113.6–114.7), 75.2% (p < 0.001; 95% CI: 53.1–97.3), 82.5% (p < 0.001; 95% CI: 76.4–88.5), 28.1% (p < 0.001; 95% CI: 14.0–42.2), and 94.2% (p < 0.001; 95% CI: 89.2–99.1). Finally, in the 14–18□Hz band, they were 76.0% (p□<□0.001; 95%□CI: 75.7–76.4), 63.6% (p□<□0.001; 95%□CI: 63.2–64.0), 117.7 (p□<□0.001; 95%□CI: 117.6–117.8), 76.2% (p□<□0.001; 95%□CI: 57.4–95.0), 82.6% (p□<□0.001; 95%□CI: 77.0–88.3), 30.3% (p□<□0.001; 95%□CI: 16.5–44.1), and 94.4% (p□<□0.001; 95%□CI: 89.4–99.3). Details of the LOO analysis are listed in Supplemental Table□4-7.

## Discussion

To our knowledge, this is the first study demonstrating that quantitative evaluation of IAP waveforms by time-frequency analysis can reliably distinguish occluded from patent states of intracranial arteries.

Arterial blood pressure waveforms consist of forward and reflected waves. The amplitude and speed of the reflected wave are positively correlated with the reflection coefficient, which reflects peripheral vessel resistance^8-10, 14^. Under occluded states, the reflected wave’s amplitude becomes larger, and it arrives earlier due to increased resistance. As a result, a distinct notch appears on the waveform, corresponding to this enhanced reflected wave; this notch is less evident or may be absent in the patent state. This morphological change in the waveform is reflected by an increase in CWT amplitude across various frequency bands. It is important to note that the width of the notch varies with heart rate (HR); when HR increases, the overall waveform narrows, causing the notch to become narrower and shift toward higher frequencies. Therefore, careful observation and comparison of changes in CWT amplitude across multiple frequency bands is essential.

Our findings showed that SAH did not significantly alter TFA with respect to intravascular states. Although increased intracranial pressure may affect IAP, it did not influence the difference in TFA between patent and occluded states. Consequently, our method can also be applied to patients with SAH.

It was demonstrated that when the effective luminal area of the catheter connected to the pressure-monitoring kit decreases, TFA is significantly reduced. Inserting additional catheters— such as distal access catheters, balloon catheters, or microcatheters—into the guiding catheter further narrows its effective lumen, presumably impairing the transmission of arterial pressure pulses and reducing waveform amplitude^17, 18^. Consequently, whenever the effective luminal area changes (e.g., through catheter insertion or removal), it is necessary to recalibrate by setting a new reference value before clinical use.

Although this effect was small, waveform trial order showed a positive correlation with TFA across all frequency bands. One possible explanation is that waveforms were acquired immediately after endovascular interventions, such as catheter and balloon manipulations in this study, thereby reflecting the progression of vasospasm triggered by these procedures. Over time, such induced spasm would presumably subside, but this was not observed in the present study, likely due to the limited measurement period.

In this study, we evaluated intravascular status based on angiographic findings. However, because angiography provides only intermittent assessments, it is difficult to precisely capture fluctuations in vascular status between imaging sessions. This represents a limitation of our study. Meanwhile, in Patient 01, the percent change in TFA showed large fluctuations crossing the cutoff (Figure 2). Because this patient experienced a thrombotic event during neuroendovascular treatment, we suspect that instability in the thrombus may have altered the intravascular status between the two angiographic examinations that confirmed occlusion. After identifying the thrombotic event, fasudil hydrochloride and ozagrel sodium were administered to dissolve the clot, and subsequent angiography revealed repeated periods of patency and occlusion. Ultimately, the clot was completely resolved, and no neurological deficits remained. These findings suggest that time-frequency analysis of IAP waveforms may enable dynamic detection of rapidly changing intravascular conditions, which are difficult to monitor with angiography alone given its intermittent observation intervals. Although angiography remains the gold standard for reliable intravascular assessment, time-frequency analysis of IAP waveforms offers an important real-time perspective. Further evaluation in a larger prospective cohort is warranted.

**Figure 2.**
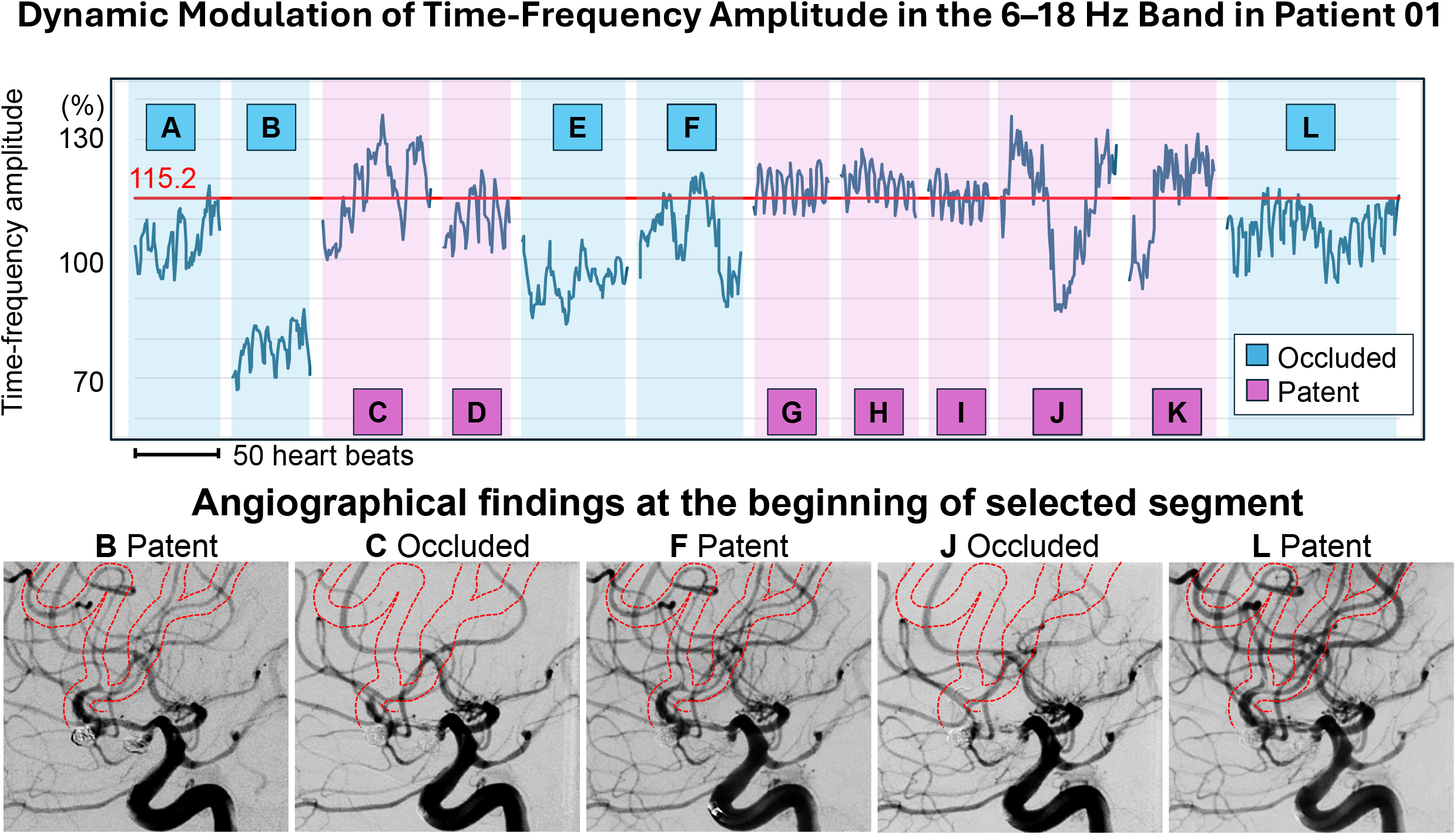
Dynamic Modulation of Time-Frequency Amplitude in the 6–18 Hz Band in Patient 01who experienced thrombosis during coil embolization for two left intracranial aneurysms. Each segment was assigned an alphabet with a background color indicating the angiographically detected intravascular states (violet: occluded; blue: patent). Although each segment was identified as either patent or occluded based on angiography, some showed considerable volatility in their percent changes, occasionally exceeding the 115.2% cutoff threshold. The lower panel displays the initial angiography of segments B, C, F, J, and L, with red dashed lines indicating the corresponding arterial structure positions (left anterior superior artery).

## Data Availability

No publicly available datasets were generated or analyzed during the present study, and the underlying data are not available.

## Data and Code Availability

No publicly available datasets were generated or analyzed during the present study, and thus the data are currently not available.

## Ethical Approval

For this retrospective study, ethical approval was obtained from the internal review boards of Iwaki City Medical Center (R6-49) and Kohnan Hospital (2024-1120-07) using an opt□out procedure.

## Acknowledgements

Study Design, Research, Data Collection, Analysis, and Manuscript Draft: Ryuzaburo Kochi, Analysis and Manuscript Draft: Eishi Asano, Data Collection: Yoshiteru Shimoda, Atsushi Kanoke, Shunsuke Omodaka, Kanako Sato, Yasuhiro Suzuki, Kuniyasu Niizuma, Hidenori Endo

## Footnote

### Nonstandard Abbreviations and Acronyms

CWT: continuous wavelet transform
IAP: intracranial arterial pressure
TFA: time-frequency amplitude

## References

1. Brinjikji, W., Murad, M. H., Lanzino, G., Cloft, H. J., & Kallmes, D. F. (2013). Endovascular treatment of intracranial aneurysms with flow diverters: a meta-analysis. Stroke, 44(2), 442–447.

2. Ries, T., Siemonsen, S., Grzyska, U., Zeumer, H., & Fiehler, J. (2009). Abciximab is a safe rescue therapy in thromboembolic events complicating cerebral aneurysm coil embolization: single center experience in 42 cases and review of the literature. Stroke, 40(5), 1750–1757.

3. Zheng, Y., Liu, Y., Leng, B., Xu, F., & Tian, Y. (2016). Periprocedural complications associated with endovascular treatment of intracranial aneurysms in 1764 cases. Journal of NeuroInterventional Surgery, 8(2), 152–157.

4. Pierot, L., Barbe, C., Nguyen, H. A., Herbreteau, D., Gauvrit, J. Y., Januel, A. C., … & White, P. (2020). Intraoperative complications of endovascular treatment of intracranial aneurysms with coiling or balloon-assisted coiling in a prospective multicenter cohort of 1088 participants: analysis of recanalization after endovascular treatment of intracranial aneurysm (ARETA) study. Radiology, 295(2), 381–389.

5. Brinjikji, W., McDonald, J. S., Kallmes, D. F., & Cloft, H. J. (2013). Rescue treatment of thromboembolic complications during endovascular treatment of cerebral aneurysms. Stroke, 44(5), 1343–1347.

6. Saver, J. L., Goyal, M., Van der Lugt, A. A. D., Menon, B. K., Majoie, C. B., Dippel, D. W., … & Hermes Collaborators. (2016). Time to treatment with endovascular thrombectomy and outcomes from ischemic stroke: a meta-analysis. Jama, 316(12), 1279–1289.

7. Adeeb, N., Griessenauer, C. J., Moore, J. M., Foreman, P. M., Shallwani, H., Motiei-Langroudi, R., … & Thomas, A. J. (2017). Ischemic stroke after treatment of intraprocedural thrombosis during stent-assisted coiling and flow diversion. Stroke, 48(4), 1098–1100.

8. Murray, W. B., & Foster, P. A. (1996). The peripheral pulse wave: information overlooked. Journal of clinical monitoring, 12, 365–377.

9. Westerhof, N., Sipkema, P., Bos, G. V. D., & Elzinga, G. (1972). Forward and backward waves in the arterial system. Cardiovascular research, 6(6), 648–656.

10. Shimoda, Y., Sonobe, S., Niizuma, K., Endo, T., Endo, H., Otomo, M., & Tominaga, T. (2021). Digital intravascular pressure wave recording during endovascular treatment reveals abnormal shunting flow in vertebral venous fistula of the vertebral artery: illustrative case. Journal of Neurosurgery: Case Lessons, 2(2).

11. De Melis, M., Morbiducci, U., Rietzschel, E. R., De Buyzere, M., Qasem, A., Van Bortel, L., … & Segers, P. (2009). Blood pressure waveform analysis by means of wavelet transform. Medical & biological engineering & computing, 47, 165–173.

12. Antonelli, L., Khamlach, R., & Ohley, W. (1994, October). Wavelet transform analysis of the arterial pressure waveform. In Proceedings of IEEE-SP International Symposium on Time-frequency and Time-Scale Analysis (pp. 568–571). IEEE.

13. Kelly, R., Hayward, C., Avolio, A., & O’rourke, M. (1989). Noninvasive determination of age-related changes in the human arterial pulse. Circulation, 80(6), 1652–1659.

14. Mynard, J. P., Kondiboyina, A., Kowalski, R., Cheung, M. M., & Smolich, J. J. (2020). Measurement, analysis and interpretation of pressure/flow waves in blood vessels. Frontiers in Physiology, 11, 1085.

15. Macdonald, R. L., & Schweizer, T. A. (2017). Spontaneous subarachnoid haemorrhage. The Lancet, 389(10069), 655–666.

16. Connolly Jr, E. S., Rabinstein, A. A., Carhuapoma, J. R., Derdeyn, C. P., Dion, J., Higashida, R. T., … & Vespa, P. (2012). Guidelines for the management of aneurysmal subarachnoid hemorrhage: a guideline for healthcare professionals from the American Heart Association/American Stroke Association. Stroke, 43(6), 1711–1737.

17. Henkes, H., Felber, S. R., Wentz, K. U., Czerwinski, F., Monstadt, H., Weber, W., & Kühne, D. (1999). Accuracy of intravascular microcatheter pressure measurements: an experimental study. The British Journal of Radiology, 72(857), 448–451.

18. Shapiro, G. G., & Krovetz, L. J. (1970). Damped and undamped frequency responses of underdamped catheter manometer systems. American Heart Journal, 80(2), 226–236.

