## Supplemental Tables 1-7 for "A Novel Use of Intracranial Arterial Pressure Waveforms to Detect Occlusive events"

Supplemental Table 1

| Pt ID | Age (range) | Sex | Occlusive event | Diagnosis | Target lesion | Procedure |
|---|---|---|---|---|---|---|
| Pt_01 | 60-69 | F | Yes | SAH | Rt . IC-PC AN | Coil embolization |
| Pt_02 | 70-79 | F | Yes | UIA | Lt. ICA lat. wall AN | BTO |
| Pt_03 | 50-59 | M | Yes | UIA | Rt . IC-PC AN | BAC |
| Pt_04 | 40-49 | M | Yes | SAH | Rt . VA AN | PAO with coils |
| Pt_05 | 80-89 | F | Yes | SAH | Rt . IC-PC AN | BAC |
| Pt_06 | 70-79 | F | Yes | UIA | Acom AN | Coil embolization |
| Pt_07 | 50-59 | F | Yes | UIA | Lt. ICA lat. wall AN | BAC |
| Pt_08 | 50-59 | F | Yes | UIA | Lt. IC_Oph | BAC |
| Pt_09 | 70-79 | F | Yes | SAH | Lt. VA AN | PAO with coils |
| Pt_10 | 40-49 | F | No | SAH | Lt. ICA lat. wall AN | Coil embolization |
| Pt_11 | 70-79 | F | No | SAH | Acom AN | Coil embolization |
| Pt_12 | 70-79 | F | No | UIA | Lt. IC_PC AN | Coil embolization |
| Pt_13 | 60-69 | M | No | UIA | Rt. MCB AN | Coil embolization |
| Pt_14 | 50-59 | M | No | UIA | Acom AN | Coil embolization |
| Pt_15 | 60-69 | F | No | UIA | Lt. IC_PC AN | Coil embolization |
| Pt_16 | 70-79 | M | No | SAH | Acom AN | Coil embolization |
| Pt_17 | 60-69 | M | Yes | SAH | Rt . IC-PC AN | BAC |
| Pt_18 | 50-59 | M | No | SAH | Acom AN | Coil embolization |
| Pt_19 | 50-59 | M | No | UIA | Rt. MCA AN | Coil embolization |
| Pt_20 | 70-79 | M | No | ICS | Lt. cervical ICA | CAS |
| Pt_21 | 40-49 | F | No | UIA | Acom AN | Coil embolization |
| Pt_22 | 60-69 | F | No | UIA | Rt . IC-PC AN | Coil embolization |
| Pt_23 | 40-49 | F | No | SAH | Lt . IC-PC | Coil embolization |
| Pt_24 | 60-69 | F | Yes | UIA | Rt. MCB AN | BAC |
| Pt_25 | 40-49 | F | Yes | SAH | Lt. ICA lat. wall AN | BAC |
| Pt_26 | 60-69 | F | No | UIA | BA AN | Coil embolization |
| Pt_27 | 50-59 | F | No | SAH | Acom AN | Coil embolization |
| Pt_28 | 40-49 | M | No | SAH | Lt. ACA AN | Coil embolization |
| Pt_29 | 60-69 | F | No | SAH | Acom AN | Coil embolization |
| Pt_30 | 70-79 | M | No | ICS | Lt. cervical ICA | CAS |
| Pt_31 | 40-49 | F | No | SAH | Lt. ICA lat. wall AN | Coil embolization |
| Pt_32 | 40-49 | F | No | SAH | BA AN | Coil embolization |
| Pt_33 | 70-79 | F | No | SAH | Lt. ICA lat. wall AN | Coil embolization |
| Pt_34 | 50-59 | F | No* | VAO | Lt. VA | PAO with coils |
| Pt_35 | 40-49 | M | Yes | SAH | Lt. VA AN | PAO with coils |
| Pt_36 | 50-59 | M | No | SAH | Rt. VA AN | Coil embolization |
| Pt_37 | 60-69 | M | Yes | UIA | Lt. ICA lat. wall AN | BAC |

Pt: patient, M: male, F: female, SAH: subarachnoid hemorrhage, UIA: unruptured intracranial aneurysm, ICS: internal carotid artery stenosis, VAO: vertebral artery occlusion, Rt: right, Lt: left, AN: aneurysm, Acom: anterior communicating artery, BA: basilar artery, ICA: internal carotid artery, IC-PC: internal carotid–posterior communicating artery, ACA: anterior cerebral artery, internal carotid ophthalmic artery, lat. wall: lateral wall, VA: vertebral artery, MCA: middle cerebral artery, MCB: middle cerebral artery bifurcation, BAC: balloon-assisted coil embolization, BTO: balloon test occlusion, CAS: carotid artery stenting, PAO: parent artery occlusion.This patient developed a left vertebral artery occlusion due to a cervical spine fracture-dislocation. Therefore, we performed PAO of the Lt. VA to prevent thrombosis during orthopedic reduction. No additional occlusive events occurred during the procedure. In this case, we only included data from the Rt. VA.

Supplemental Table 2

| Pt ID | Sampled artery | Segmet number | Intravascular states | Occlusion point | Occlusive etiology | Effective luminal area of the guiding catheter ($mm^2$) |
|---|---|---|---|---|---|---|
| Pt_01 | Rt. ICA | Segment_01 | Patent | na | na | 3.30 |
| | | Segment_02 | Patent | na | na | 1.76 |
| | | Segment_03 | Patent | na | na | 1.76 |
| | | Segment_04 | Occlusive | MCA bifurcation | Clot | 1.76 |
| | | Segment_05 | Occlusive | MCA bifurcation | Clot | 1.76 |
| | | Segment_06 | Patent | na | na | 1.76 |
| | | Segment_07 | Occlusive | MCA bifurcation | Clot | 1.76 |
| | | Segment_08 | Occlusive | MCA bifurcation | Clot | 1.76 |
| | | Segment_09 | Occlusive | MCA bifurcation | Clot | 1.76 |
| | | Segment_10 | Occlusive | MCA bifurcation | Clot | 1.76 |
| | | Segment_11 | Occlusive | MCA bifurcation | Clot | 1.76 |
| | | Segment_12 | Occlusive | MCA bifurcation | Clot | 1.76 |
| | | Segment_13 | Patent | na | na | 1.76 |
| Pt_02 | Lt. ICA | Segment_01 | Patent | na | na | 1.22 |
| | | Segment_02 | Occlusive | Petrus segment | Balloon (3.4Fr MASAMUNE) | 1.22 |
| | Lt. VA | Segment_01 | Patent | na | na | 0.87 |
| | Rt. ICA | Segment_01 | Patent | na | na | 0.87 |
| Pt_03 | Rt. ICA | Segment_01 | Patent | na | na | 2.77 |
| | | Segment_02 | Occlusive | IC-PC | Balloon (SHORYU HR 4x7) | 2.77 |
| Pt_04 | Rt. VA | Segment_01 | Patent | na | na | 2.54 |
| | | Segment_02 | Occlusive | V4 segment | Coil | 2.54 |
| | | Segment_03 | Occlusive | V4 segment | Coil | 2.54 |
| Pt_05 | Rt. ICA | Segment_01 | Patent | na | na | 4.08 |
| | | Segment_02 | Patent | na | na | 2.77 |
| | | Segment_03 | Occlusive | IC-PC segment | Balloon (SHORYU HR 4x7) | 2.77 |
| | | Segment_04 | Occlusive | IC-PC segment | Balloon (SHORYU HR 4x7) | 2.77 |
| | | Segment_05 | Patent | na | na | 2.77 |
| | | Segment_06 | Patent | na | na | 4.08 |
| Pt_06 | Lt. ICA | Segment_01 | Patent | na | na | 4.08 |
| | | Segment_02 | Occlusive | A1 | Clot | 4.08 |
| Pt_07 | Lt. ICA | Segment_01 | Patent | na | na | 4.08 |
| | | Segment_02 | Occlusive | na | na | 1.59 |
| | | Segment_03 | Occlusive | Supraclinoid segmet | Balloon (TransForm 7x7) | 1.59 |
| | | Segment_04 | Patent | Supraclinoid segmet | Balloon (TransForm 7x7) | 1.59 |
| | | Segment_05 | Patent | na | na | 1.59 |
| | | Segment_06 | Patent | na | na | 4.08 |
| Pt_08 | Lt. ICA | Segment_01 | Patent | na | na | 2.61 |
| | | Segment_02 | Occlusive | IC_Ophthalmic segment | Balloon (Scepter XC 4x11) | 2.61 |
| | | Segment_03 | Patent | na | na | 2.61 |
| | | Segment_04 | Occlusive | IC_Ophthalmic segment | Balloon (Scepter XC 4x11) | 2.61 |
| | | Segment_05 | Patent | na | na | 2.61 |

| | | | | | | |
|---|---|---|---|---|---|---|
| Pt_09 | Lt. VA | Segment_01 | Patent | na | na | 2.11 |
| | | Segment_02 | Patent | na | na | 2.11 |
| | | Segment_03 | Patent | na | na | 2.11 |
| | | Segment_04 | Patent | na | na | 2.11 |
| | | Segment_05 | Patent | na | na | 2.11 |
| | | Segment_06 | Patent | na | na | 2.11 |
| | | Segment_07 | Patent | na | na | 2.11 |
| | | Segment_08 | Occlusive | V4 | Coil | 2.11 |
| | | Segment_09 | Occlusive | V4 | Coil | 2.11 |
| | | Segment_10 | Occlusive | V4 | Coil | 2.11 |
| | | Segment_11 | Occlusive | V4 | Coil | 2.11 |
| | | Segment_12 | Occlusive | V4 | Coil | 2.11 |
| | | Segment_13 | Occlusive | V4 | Coil | 2.11 |
| | Rt. ICA | Segment_01 | Patent | na | na | 0.87 |
| | Lt. ICA | Segment_01 | Patent | na | na | 0.87 |
| Pt_10 | Lt. ICA | Segment_01 | Patent | na | na | 4.08 |
| Pt_11 | Lt. ICA | Segment_01 | Patent | na | na | 4.08 |
| Pt_12 | Lt. ICA | Segment_01 | Patent | na | na | 4.08 |
| Pt_13 | Rt. ICA | Segment_01 | Patent | na | na | 4.08 |
| Pt_14 | Lt. ICA | Segment_01 | Patent | na | na | 4.08 |
| Pt_15 | Lt. ICA | Segment_01 | Patent | na | na | 3.18 |
| | | Segment_02 | Patent | na | na | 3.18 |
| | | Segment_03 | Patent | na | na | 4.08 |
| Pt_16 | Rt. ICA | Segment_01 | Patent | na | na | 3.30 |
| Pt_17 | Rt. ICA | Segment_01 | Patent | na | na | 3.00 |
| | | Segment_02 | Occlusive | IC_PC | Balloon (Scepter XC 4x11) | 3.00 |
| Pt_18 | Lt. ICA | Segment_01 | Patent | na | na | 1.76 |
| Pt_19 | Rt. ICA | Segment_01 | Patent | na | na | 3.30 |
| Pt_20 | Lt. VA | Segment_01 | Patent | na | na | 0.66 |
| | | Segment_02 | Patent | na | na | 0.66 |
| Pt_21 | Lt. ICA | Segment_01 | Patent | na | na | 1.76 |
| | | Segment_02 | Patent | na | na | 3.30 |
| Pt_22 | Rt. ICA | Segment_01 | Patent | na | na | 5.07 |
| | | Segment_02 | Patent | na | na | 5.07 |
| Pt_23 | Lt. ICA | Segment_01 | Patent | na | na | 2.54 |

| | | | | | | |
|---|---|---|---|---|---|---|
| Pt_24 | Rt. ICA | Segment_01 | Patent | na | na | 5.07 |
| | | Segment_02 | Patent | na | na | 1.60 |
| | | Segment_03 | Occlusive | MCA bifurcation | Balloon (Scepter XC 4x11) | 1.60 |
| | | Segment_04 | Occlusive | MCA bifurcation | Balloon (Scepter XC 4x11) | 1.60 |
| | | Segment_05 | Occlusive | MCA bifurcation | Balloon (Scepter XC 4x11) | 1.60 |
| | | Segment_06 | Patent | na | na | 5.07 |
| Pt_25 | Lt. ICA | Segment_01 | Patent | na | na | 2.54 |
| | | Segment_02 | Patent | na | na | 2.54 |
| | | Segment_03 | Patent | na | na | 1.33 |
| | | Segment_04 | Occlusive | Supraclinoid segmet | Balloon (TransForm 4x10) | 1.33 |
| | | Segment_05 | Patent | na | na | 1.33 |
| | | Segment_06 | Occlusive | Supraclinoid segmet | Balloon (TransForm 4x10) | 1.33 |
| | | Segment_07 | Patent | na | na | 1.33 |
| | | Segment_08 | Occlusive | Supraclinoid segmet | Balloon (TransForm 4x10) | 1.33 |
| | | Segment_09 | Patent | na | na | 1.33 |
| | | Segment_10 | Patent | na | na | 2.54 |
| Pt_26 | Lt. VA | Segment_01 | Patent | na | na | 1.99 |
| | | Segment_02 | Patent | na | na | 1.99 |
| | | Segment_03 | Patent | na | na | 2.54 |
| | Rt. VA | Segment_01 | Patent | na | na | 1.12 |
| | | Segment_02 | Patent | na | na | 1.70 |
| Pt_27 | Lt. ICA | Segment_01 | Patent | na | na | 3.30 |
| Pt_28 | Lt. ICA | Segment_01 | Patent | na | na | 5.07 |
| | | Segment_02 | Patent | na | na | 5.07 |
| Pt_29 | Lt. ICA | Segment_01 | Patent | na | na | 5.07 |
| | | Segment_02 | Patent | na | na | 5.07 |
| | | Segment_03 | Patent | na | na | 5.07 |
| | | Segment_05 | Patent | na | na | 3.53 |
| | | Segment_06 | Patent | na | na | 5.07 |
| Pt_30 | Lt. VA | Segment_01 | Patent | na | na | 0.87 |
| Pt_31 | Lt. ICA | Segment_01 | Patent | na | na | 2.54 |
| Pt_32 | Rt. VA | Segment_01 | Patent | na | na | 2.54 |
| Pt_33 | Lt. ICA | Segment_01 | Patent | na | na | 4.08 |
| | | Segment_02 | Patent | na | na | 4.08 |
| Pt_34 | Rt. VA | Segment_01 | Patent | na | na | 0.66 |
| Pt_35 | Lt. VA | Segment_01 | Patent | na | na | 2.54 |
| | | Segment_02 | Patent | na | na | 2.04 |
| | | Segment_03 | Patent | na | na | 2.04 |
| | | Segment_04 | Patent | na | na | 2.04 |
| | | Segment_05 | Occlusive | V4 segment | Coil | 2.04 |
| | Rt. VA | Segment_01 | Patent | na | na | 0.66 |
| Pt_36 | Lt. VA | Segment_01 | Patent | na | na | 0.66 |
| Pt_37 | Lt. ICA | Segment_01 | Patent | na | na | 4.08 |
| | | Segment_02 | Patent | na | na | 2.61 |
| | | Segment_03 | Occlusive | Supraclinoid segmet | Balloon (Scepter XC 4x11) | 2.61 |
| | | Segment_04 | Occlusive | Supraclinoid segmet | Balloon (Scepter XC 4x11) | 2.61 |
| | | Segment_05 | Occlusive | Supraclinoid segmet | Balloon (Scepter XC 4x11) | 2.61 |
| | | Segment_06 | Patent | na | na | 4.08 |

Pt: patient, Rt: right, Lt: left, ICA: internal carotid artery, VA: vertebral artery DAC: distal access catheter, Scepter XC: Scepter XC microballoon (MicroVention, Tustin, CA), Transform: Transform (Stryker, Fremont, CA, USA), SHORYU HR: SHORYU HR (Kaneka, Osaka, Japan)

Supplemental Table 3

**Occluded states**

| Frequency bands (Hz) | Estimate | Standard error | Degrees of freedom | t-value | p-value | 95% Confidence Interval Lower | 95% Confidence Interval Upper |
|---|---|---|---|---|---|---|---|
| 6-10 | 0.017 | 0 | 6414.054 | 55.589 | <.001 | 0.016 | 0.017 |
| 10-14 | 0.011 | 0 | 6424.174 | 49.37 | <.001 | 0.011 | 0.011 |
| 14-18 | 0.002 | 0 | 6427.641 | 20.764 | <.001 | 0.002 | 0.003 |
| 6-18 | 0.03 | 0.001 | 6416.066 | 55.413 | <.001 | 0.029 | 0.031 |

**Waveform trial order**

| Frequency bands (Hz) | Estimate | Standard error | Degrees of freedom | t-value | p-value | 95% Confidence Interval Lower | 95% Confidence Interval Upper |
|---|---|---|---|---|---|---|---|
| 6-10 | $1.49\times10^{-5}$ | $3.08\times10^{-6}$ | 6406.642 | 4.819 | <.001 | $8.81\times10^{-6}$ | $2.09\times10^{-5}$ |
| 10-14 | $1.05\times10^{-5}$ | $2.30\times10^{-6}$ | 6413.793 | 4.571 | <.001 | $6.02\times10^{-6}$ | $1.51\times10^{-5}$ |
| 14-18 | $6.20\times10^{-6}$ | $1.22\times10^{-6}$ | 6417.561 | 5.071 | <.001 | $3.81\times10^{-6}$ | $8.60\times10^{-6}$ |
| 6-18 | $3.16\times10^{-5}$ | $5.60\times10^{-6}$ | 6407.969 | 5.636 | <.001 | $2.06\times10^{-5}$ | $4.26\times10^{-5}$ |

**The effective luminal area of the catheter connected to the pressure monitoring kit**

| Frequency bands (Hz) | Estimate | Standard error | Degrees of freedom | t-value | p-value | 95% Confidence Interval Lower | 95% Confidence Interval Upper |
|---|---|---|---|---|---|---|---|
| 6-10 | 0.006 | 0 | 6410.537 | 30.893 | <.001 | 0.006 | 0.007 |
| 10-14 | 0.005 | 0 | 6249.821 | 30.069 | <.001 | 0.004 | 0.005 |
| 14-18 | 0.003 | $8.26\times10^{-5}$ | 6331.624 | 38.946 | <.001 | 0.003 | 0.003 |
| 6-18 | 0.014 | 0 | 6394.518 | 37.851 | <.001 | 0.014 | 0.015 |

Supplemental Table 4: Result of 6-18 Hz bands

| Left out artery | AUROC | AUPRC | Youden Index | Best Cutoff | Sensitivity (%) | Specificity (%) | PPV (%) | NPV (%) |
|---|---|---|---|---|---|---|---|---|
| Artery_01 | 0.93 | 0.82 | 0.77 | 115.18 | 53.4 | 97.9 | 97.2 | 60.7 |
| Artery_02 | 0.88 | 0.77 | 0.67 | 114.06 | 95.1 | 100 | 100 | 90.9 |
| Artery_03 | 0.88 | 0.77 | 0.67 | 114.06 | | 100 | | 100 |
| Artery_04 | 0.88 | 0.77 | 0.67 | 114.06 | | 100 | | 100 |
| Artery_05 | 0.88 | 0.77 | 0.67 | 114.06 | 100 | 100 | 100 | 100 |
| Artery_06 | 0.87 | 0.75 | 0.66 | 114.06 | 100 | 97.3 | 97.8 | 100 |
| Artery_07 | 0.88 | 0.74 | 0.67 | 113.38 | 100 | 53.5 | 61.1 | 100 |
| Artery_08 | 0.88 | 0.77 | 0.67 | 114.06 | 86.2 | 89.3 | 94.3 | 75.8 |
| Artery_09 | 0.88 | 0.77 | 0.66 | 113.38 | 100 | 94.5 | 92.2 | 100 |
| Artery_10 | 0.90 | 0.79 | 0.69 | 113.38 | 100 | 23.4 | 48.8 | 100 |
| Artery_11 | 0.87 | 0.76 | 0.64 | 113.38 | 100 | 100 | 100 | 100 |
| Artery_12 | 0.88 | 0.77 | 0.67 | 114.06 | | 100 | | 100 |
| Artery_13 | 0.88 | 0.77 | 0.67 | 114.06 | | 50.0 | 0 | 100 |
| Artery_14 | 0.88 | 0.77 | 0.67 | 114.06 | | 100 | | 100 |
| Artery_15 | 0.88 | 0.77 | 0.67 | 114.06 | | 73.3 | 0 | 100 |
| Artery_16 | 0.88 | 0.78 | 0.68 | 114.06 | | 21.4 | 0 | 100 |
| Artery_17 | 0.88 | 0.77 | 0.67 | 114.06 | | 100 | | 100 |
| Artery_18 | 0.88 | 0.77 | 0.67 | 114.06 | | 100 | | 100 |
| Artery_19 | 0.88 | 0.78 | 0.68 | 114.06 | | 62.4 | 0 | 100 |
| Artery_20 | 0.88 | 0.77 | 0.67 | 114.06 | | 100 | | 100 |
| Artery_21 | 0.89 | 0.78 | 0.68 | 114.06 | 14.8 | 100 | 100 | 48.9 |
| Artery_22 | 0.88 | 0.77 | 0.67 | 114.06 | | 94.4 | 0 | 100 |
| Artery_23 | 0.88 | 0.77 | 0.67 | 114.06 | | 78.6 | 0 | 100 |
| Artery_24 | 0.88 | 0.77 | 0.67 | 114.06 | | 100 | | 100 |
| Artery_25 | 0.88 | 0.77 | 0.67 | 114.06 | | 97.3 | 0 | 100 |
| Artery_26 | 0.88 | 0.77 | 0.67 | 114.06 | | 100 | | 100 |
| Artery_27 | 0.88 | 0.77 | 0.67 | 114.06 | | 100 | | 100 |
| Artery_28 | 0.87 | 0.75 | 0.66 | 113.38 | 100 | 90.0 | 82.6 | 100 |
| Artery_29 | 0.88 | 0.79 | 0.68 | 114.06 | 100 | 62.5 | 33.9 | 100 |
| Artery_30 | 0.88 | 0.77 | 0.67 | 114.06 | | 99.1 | 0 | 100 |
| Artery_31 | 0.88 | 0.77 | 0.67 | 114.06 | | 94.7 | 0 | 100 |
| Artery_32 | 0.88 | 0.77 | 0.67 | 114.06 | | 95.0 | 0 | 100 |
| Artery_33 | 0.88 | 0.77 | 0.67 | 114.06 | | 98.0 | 0 | 100 |
| Artery_34 | 0.88 | 0.77 | 0.66 | 114.06 | | 99.1 | 0 | 100 |
| Artery_35 | 0.88 | 0.77 | 0.67 | 114.06 | | 92.6 | 0 | 100 |
| Artery_36 | 0.88 | 0.78 | 0.67 | 111.22 | | 68.9 | 0 | 100 |
| Artery_37 | 0.88 | 0.77 | 0.66 | 114.06 | | 99.5 | 0 | 100 |
| Artery_38 | 0.88 | 0.77 | 0.67 | 114.06 | | 100 | | 100 |
| Artery_39 | 0.88 | 0.77 | 0.67 | 114.06 | | 96.8 | 0 | 100 |
| Artery_40 | 0.88 | 0.78 | 0.68 | 114.06 | 100 | 55.2 | 13.9 | 100 |
| Artery_41 | 0.88 | 0.77 | 0.67 | 114.06 | | 100 | | 100 |
| Artery_42 | 0.88 | 0.77 | 0.67 | 114.06 | | 90.3 | 0 | 100 |
| Artery_43 | 0.88 | 0.76 | 0.66 | 113.38 | 89.8 | 87.1 | 96.0 | 71.1 |

AUROC: area under the receiver operating characteristic curve, AUPRC: area under the precision–recall curve, PPV: positive predictive value, NPV: negative predictive value

Supplemental Table 5: Result of 6-10 Hz bands

| Left out artery | AUROC | AUPRC | Youden Index | Best Cutoff | Sensitivity (%) | Specificity (%) | PPV (%) | NPV (%) |
|---|---|---|---|---|---|---|---|---|
| Artery_01 | 0.87 | 0.81 | 0.69 | 124.35 | 25.8 | 97.9 | 94.3 | 49.2 |
| Artery_02 | 0.84 | 0.76 | 0.54 | 113.58 | 100 | 100 | 100 | 100 |
| Artery_03 | 0.84 | 0.76 | 0.55 | 113.58 | | 63.6 | 0 | 100 |
| Artery_04 | 0.84 | 0.76 | 0.54 | 113.58 | | 94.6 | 0 | 100 |
| Artery_05 | 0.84 | 0.76 | 0.54 | 113.58 | 100 | 100 | 100 | 100 |
| Artery_06 | 0.83 | 0.75 | 0.54 | 113.58 | 85.7 | 93.3 | 94.0 | 84.3 |
| Artery_07 | 0.83 | 0.73 | 0.54 | 113.58 | 100 | 53.1 | 60.9 | 100 |
| Artery_08 | 0.85 | 0.77 | 0.57 | 113.58 | 3.4 | 78.6 | 25.0 | 28.2 |
| Artery_09 | 0.87 | 0.78 | 0.58 | 113.57 | 1.2 | 89.0 | 6.7 | 57.9 |
| Artery_10 | 0.83 | 0.74 | 0.54 | 113.58 | 100 | 43.4 | 56.3 | 100 |
| Artery_11 | 0.81 | 0.71 | 0.50 | 113.58 | 100.0 | 99.6 | 99.6 | 100 |
| Artery_12 | 0.84 | 0.76 | 0.54 | 113.58 | | 100 | | 100 |
| Artery_13 | 0.84 | 0.76 | 0.55 | 113.57 | | 55.0 | 0 | 100 |
| Artery_14 | 0.84 | 0.76 | 0.54 | 113.58 | | 100 | | 100 |
| Artery_15 | 0.84 | 0.76 | 0.55 | 113.58 | | 60.0 | 0 | 100 |
| Artery_16 | 0.84 | 0.76 | 0.55 | 113.58 | | 50.0 | 0 | 100 |
| Artery_17 | 0.84 | 0.76 | 0.54 | 113.58 | | 100 | | 100 |
| Artery_18 | 0.84 | 0.76 | 0.54 | 113.58 | | 100 | | 100 |
| Artery_19 | 0.84 | 0.77 | 0.55 | 116.91 | | 61.5 | 0 | 100 |
| Artery_20 | 0.84 | 0.76 | 0.54 | 113.58 | | 100 | | 100 |
| Artery_21 | 0.84 | 0.76 | 0.55 | 113.58 | 48.1 | 100 | 100 | 61.1 |
| Artery_22 | 0.84 | 0.76 | 0.54 | 113.58 | | 77.8 | 0 | 100 |
| Artery_23 | 0.84 | 0.76 | 0.55 | 113.58 | | 73.2 | 0 | 100 |
| Artery_24 | 0.84 | 0.76 | 0.54 | 113.58 | | 96.4 | 0 | 100 |
| Artery_25 | 0.84 | 0.76 | 0.54 | 113.58 | | 100 | | 100 |
| Artery_26 | 0.84 | 0.76 | 0.54 | 116.91 | | 100 | | 100 |
| Artery_27 | 0.84 | 0.76 | 0.54 | 113.58 | | 100 | | 100 |
| Artery_28 | 0.83 | 0.74 | 0.53 | 113.58 | 100 | 79.6 | 69.9 | 100 |
| Artery_29 | 0.84 | 0.77 | 0.54 | 113.57 | 100 | 67.2 | 37.0 | 100 |
| Artery_30 | 0.84 | 0.76 | 0.54 | 113.58 | | 92.8 | 0 | 100 |
| Artery_31 | 0.84 | 0.76 | 0.54 | 113.58 | | 97.3 | 0 | 100 |
| Artery_32 | 0.84 | 0.76 | 0.54 | 113.58 | | 95.0 | 0 | 100 |
| Artery_33 | 0.84 | 0.77 | 0.55 | 116.72 | | 43 | 0 | 100 |
| Artery_34 | 0.84 | 0.76 | 0.54 | 113.58 | | 88.0 | 0 | 100 |
| Artery_35 | 0.84 | 0.76 | 0.54 | 113.58 | | 88.9 | 0 | 100 |
| Artery_36 | 0.84 | 0.78 | 0.57 | 113.58 | | 50.7 | 0 | 100 |
| Artery_37 | 0.84 | 0.76 | 0.54 | 116.91 | | 94.4 | 0 | 100 |
| Artery_38 | 0.84 | 0.76 | 0.54 | 113.58 | | 100 | | 100 |
| Artery_39 | 0.84 | 0.76 | 0.54 | 113.58 | | 93.5 | 0 | 100 |
| Artery_40 | 0.84 | 0.76 | 0.55 | 113.58 | 100 | 61.3 | 15.7 | 100 |
| Artery_41 | 0.84 | 0.76 | 0.54 | 113.58 | | 81.1 | 0 | 100 |
| Artery_42 | 0.84 | 0.77 | 0.55 | 113.58 | | 39.8 | 0 | 100 |
| Artery_43 | 0.84 | 0.75 | 0.53 | 113.58 | 88.4 | 87.1 | 96.0 | 68.4 |

AUROC: area under the receiver operating characteristic curve, AUPRC: area under the precision–recall curve, PPV: positive predictive value, NPV: negative predictive value

Supplemental Table 6: Result of 10-14 Hz bands

| Left out artery | AUROC | AUPRC | Youden Index | Best Cutoff | Sensitivity (%) | Specificity (%) | PPV (%) | NPV (%) |
|---|---|---|---|---|---|---|---|---|
| Artery_01 | 0.89 | 0.74 | 0.63 | 110.36 | 53.9 | 97.9 | 97.2 | 60.9 |
| Artery_02 | 0.84 | 0.71 | 0.58 | 109.00 | 2.4 | 100 | 100 | 33.3 |
| Artery_03 | 0.84 | 0.71 | 0.56 | 107.13 | | 95.5 | 0 | 100 |
| Artery_04 | 0.84 | 0.71 | 0.56 | 107.13 | | 100 | | 100 |
| Artery_05 | 0.84 | 0.70 | 0.56 | 107.13 | 100 | 84.2 | 78.6 | 100 |
| Artery_06 | 0.83 | 0.68 | 0.56 | 107.13 | 100 | 69.3 | 79.8 | 100 |
| Artery_07 | 0.83 | 0.69 | 0.55 | 107.13 | 100 | 59.8 | 64.5 | 100 |
| Artery_08 | 0.84 | 0.71 | 0.57 | 109.59 | 46.6 | 89.3 | 90.0 | 44.6 |
| Artery_09 | 0.83 | 0.70 | 0.55 | 107.13 | 100 | 79.5 | 76.1 | 100 |
| Artery_10 | 0.85 | 0.74 | 0.58 | 107.13 | 100 | 21.1 | 48.1 | 100 |
| Artery_11 | 0.84 | 0.71 | 0.57 | 110.78 | 53.8 | 100 | 100 | 67.2 |
| Artery_12 | 0.84 | 0.71 | 0.56 | 107.13 | | 81.8 | 0 | 100 |
| Artery_13 | 0.84 | 0.71 | 0.57 | 107.13 | | 25.0 | 0 | 100 |
| Artery_14 | 0.84 | 0.71 | 0.56 | 107.13 | | 100 | | 100 |
| Artery_15 | 0.84 | 0.71 | 0.56 | 107.13 | | 66.7 | 0 | 100 |
| Artery_16 | 0.84 | 0.71 | 0.57 | 107.13 | | 12.5 | 0 | 100 |
| Artery_17 | 0.84 | 0.71 | 0.56 | 107.13 | | 100 | | 100 |
| Artery_18 | 0.84 | 0.71 | 0.56 | 107.13 | | 100 | | 100 |
| Artery_19 | 0.84 | 0.71 | 0.57 | 107.13 | | 56.0 | 0 | 100 |
| Artery_20 | 0.84 | 0.71 | 0.56 | 107.13 | | 100 | | 100 |
| Artery_21 | 0.85 | 0.71 | 0.57 | 107.13 | 7.4 | 100 | 100 | 46.8 |
| Artery_22 | 0.84 | 0.71 | 0.56 | 107.13 | | 88.9 | 0 | 100 |
| Artery_23 | 0.84 | 0.71 | 0.57 | 107.13 | | 60.7 | 0 | 100 |
| Artery_24 | 0.84 | 0.71 | 0.56 | 107.13 | | 100 | | 100 |
| Artery_25 | 0.84 | 0.71 | 0.56 | 107.13 | | 73.0 | 0 | 100 |
| Artery_26 | 0.84 | 0.71 | 0.56 | 107.13 | | 91.4 | 0 | 100 |
| Artery_27 | 0.84 | 0.71 | 0.56 | 107.13 | | 100 | | 100 |
| Artery_28 | 0.83 | 0.68 | 0.55 | 107.13 | 100 | 78.6 | 68.8 | 100 |
| Artery_29 | 0.84 | 0.72 | 0.57 | 107.13 | 100 | 57.3 | 31.1 | 100 |
| Artery_30 | 0.84 | 0.71 | 0.56 | 107.13 | | 78.4 | 0 | 100 |
| Artery_31 | 0.84 | 0.71 | 0.56 | 107.13 | | 86.7 | 0 | 100 |
| Artery_32 | 0.84 | 0.71 | 0.56 | 107.13 | | 95.0 | 0 | 100 |
| Artery_33 | 0.84 | 0.71 | 0.56 | 107.13 | | 100 | | 100 |
| Artery_34 | 0.84 | 0.71 | 0.56 | 107.13 | | 81.8 | 0 | 100 |
| Artery_35 | 0.84 | 0.71 | 0.57 | 107.13 | | 37.0 | 0 | 100 |
| Artery_36 | 0.84 | 0.71 | 0.58 | 107.13 | | 61.7 | 0 | 100 |
| Artery_37 | 0.83 | 0.71 | 0.55 | 109.00 | | 97.0 | 0 | 100 |
| Artery_38 | 0.84 | 0.71 | 0.56 | 107.13 | | 98.4 | 0 | 100 |
| Artery_39 | 0.84 | 0.71 | 0.56 | 107.13 | | 90.3 | 0 | 100 |
| Artery_40 | 0.84 | 0.72 | 0.58 | 107.13 | 100 | 35.6 | 10.1 | 100 |
| Artery_41 | 0.84 | 0.71 | 0.56 | 107.13 | | 96.2 | 0 | 100 |
| Artery_42 | 0.84 | 0.71 | 0.56 | 107.13 | | 88.2 | 0 | 100 |
| Artery_43 | 0.83 | 0.68 | 0.54 | 107.13 | 98.6 | 67.7 | 91.4 | 93.3 |

AUROC: area under the receiver operating characteristic curve, AUPRC: area under the precision–recall curve, PPV: positive predictive value, NPV: negative predictive value

Supplemental Table 7: Result of 14-18 Hz bands

| Left out artery | AUROC | AUPRC | Youden Index | Best Cutoff | Sensitivity (%) | Specificity (%) | PPV (%) | NPV (%) |
|---|---|---|---|---|---|---|---|---|
| Artery_01 | 0.89 | 0.74 | 0.63 | 110.36 | 53.9 | 97.9 | 97.2 | 60.9 |
| Artery_02 | 0.84 | 0.71 | 0.58 | 109.00 | 2.4 | 100 | 100 | 33.3 |
| Artery_03 | 0.84 | 0.71 | 0.56 | 107.13 | | 95.5 | 0 | 100 |
| Artery_04 | 0.84 | 0.71 | 0.56 | 107.13 | | 100 | | 100 |
| Artery_05 | 0.84 | 0.70 | 0.56 | 107.13 | 100 | 84.2 | 78.6 | 100 |
| Artery_06 | 0.83 | 0.68 | 0.56 | 107.13 | 100 | 69.3 | 79.8 | 100 |
| Artery_07 | 0.83 | 0.69 | 0.55 | 107.13 | 100 | 59.8 | 64.5 | 100 |
| Artery_08 | 0.84 | 0.71 | 0.57 | 109.59 | 46.6 | 89.3 | 90.0 | 44.6 |
| Artery_09 | 0.83 | 0.70 | 0.55 | 107.13 | 100 | 79.5 | 76.1 | 100 |
| Artery_10 | 0.85 | 0.74 | 0.58 | 107.13 | 100 | 21.1 | 48.1 | 100 |
| Artery_11 | 0.84 | 0.71 | 0.57 | 110.78 | 53.8 | 100 | 100 | 67.2 |
| Artery_12 | 0.84 | 0.71 | 0.56 | 107.13 | | 81.8 | 0 | 100 |
| Artery_13 | 0.84 | 0.71 | 0.57 | 107.13 | | 25.0 | 0 | 100 |
| Artery_14 | 0.84 | 0.71 | 0.56 | 107.13 | | 100 | | 100 |
| Artery_15 | 0.84 | 0.71 | 0.56 | 107.13 | | 66.7 | 0 | 100 |
| Artery_16 | 0.84 | 0.71 | 0.57 | 107.13 | | 12.5 | 0 | 100 |
| Artery_17 | 0.84 | 0.71 | 0.56 | 107.13 | | 100 | | 100 |
| Artery_18 | 0.84 | 0.71 | 0.56 | 107.13 | | 100 | | 100 |
| Artery_19 | 0.84 | 0.71 | 0.57 | 107.13 | | 56.0 | 0 | 100 |
| Artery_20 | 0.84 | 0.71 | 0.56 | 107.13 | | 100 | | 100 |
| Artery_21 | 0.85 | 0.71 | 0.57 | 107.13 | 7.4 | 100 | 100 | 46.8 |
| Artery_22 | 0.84 | 0.71 | 0.56 | 107.13 | | 88.9 | 0 | 100 |
| Artery_23 | 0.84 | 0.71 | 0.57 | 107.13 | | 60.7 | 0 | 100 |
| Artery_24 | 0.84 | 0.71 | 0.56 | 107.13 | | 100 | | 100 |
| Artery_25 | 0.84 | 0.71 | 0.56 | 107.13 | | 73.0 | 0 | 100 |
| Artery_26 | 0.84 | 0.71 | 0.56 | 107.13 | | 91.4 | 0 | 100 |
| Artery_27 | 0.84 | 0.71 | 0.56 | 107.13 | | 100 | | 100 |
| Artery_28 | 0.83 | 0.68 | 0.55 | 107.13 | 100 | 78.6 | 68.8 | 100 |
| Artery_29 | 0.84 | 0.72 | 0.57 | 107.13 | 100 | 57.3 | 31.1 | 100 |
| Artery_30 | 0.84 | 0.71 | 0.56 | 107.13 | | 78.4 | 0 | 100 |
| Artery_31 | 0.84 | 0.71 | 0.56 | 107.13 | | 86.7 | 0 | 100 |
| Artery_32 | 0.84 | 0.71 | 0.56 | 107.13 | | 95.0 | 0 | 100 |
| Artery_33 | 0.84 | 0.71 | 0.56 | 107.13 | | 100 | | 100 |
| Artery_34 | 0.84 | 0.71 | 0.56 | 107.13 | | 81.8 | 0 | 100 |
| Artery_35 | 0.84 | 0.71 | 0.57 | 107.13 | | 37.0 | 0 | 100 |
| Artery_36 | 0.84 | 0.71 | 0.58 | 107.13 | | 61.7 | 0 | 100 |
| Artery_37 | 0.83 | 0.71 | 0.55 | 109.00 | | 97.0 | 0 | 100 |
| Artery_38 | 0.84 | 0.71 | 0.56 | 107.13 | | 98.4 | 0 | 100 |
| Artery_39 | 0.84 | 0.71 | 0.56 | 107.13 | | 90.3 | 0 | 100 |
| Artery_40 | 0.84 | 0.72 | 0.58 | 107.13 | 100 | 35.6 | 10.1 | 100 |
| Artery_41 | 0.84 | 0.71 | 0.56 | 107.13 | | 96.2 | 0 | 100 |
| Artery_42 | 0.84 | 0.71 | 0.56 | 107.13 | | 88.2 | 0 | 100 |
| Artery_43 | 0.83 | 0.68 | 0.54 | 107.13 | 98.6 | 67.7 | 91.4 | 93.3 |

AUROC: area under the receiver operating characteristic curve, AUPRC: area under the precision–recall curve, PPV: positive predictive value, NPV: negative predictive value
